# SEVA: An externally driven framework for reproducing COVID-19 mortality waves without transmission feedback

**DOI:** 10.64898/2026.01.30.26345245

**Authors:** Kim Varming

## Abstract

Understanding the mechanisms underlying epidemic waveform structure remains a central problem in infectious disease epidemiology. During the first COVID-19 wave, epidemic time series across regions displayed markedly different temporal patterns, including sharply peaked curves with rapid decline and more prolonged plateau-like trajectories.

Here we present a parsimonious dynamical framework for describing epidemic waveforms at the level of observable endpoints. In the SEVA (Seasonal/Environmental Viral Activity) formulation, epidemic incidence arises from the interaction between a time-varying activity function, interpreted as a population-level hazard, and depletion of a finite vulnerable population. With activation timing and steepness held constant, variation in activity intensity governs the resulting waveform dynamics.

The framework is applied to hospitalization and mortality data from European countries and U.S. states during spring 2020. Using a common activation profile, the model reproduces both sharply peaked and plateau-like epidemic regimes across regions without modification of the underlying dynamical structure. While interpersonal transmission undoubtedly occurs, the observed epidemic waveforms are not readily explained by transmission feedback alone.

A key empirical observation is that normalized epidemic trajectories exhibit closely similar temporal structure across regions with widely different mortality burdens. Within the proposed framework, this behaviour can be understood as arising from the interaction between a shared temporal activation profile and region-specific variation in activity intensity.

These results show that diverse epidemic waveform regimes can be captured within a simple dynamical framework operating at the level of observable endpoints.

The SEVA framework therefore provides a testable hypothesis for the dynamical origin of epidemic waveforms that can be evaluated across different pathogens and epidemiological settings.

## 1. Introduction

Mathematical models play a central role in describing the dynamical behaviour of infectious disease epidemics at the population level. In many classical formulations, epidemic growth and decline arise from interaction-driven feedback between susceptible and infectious individuals. In such SIR-type frameworks, population-level epidemic waveforms emerge from nonlinear transmission processes coupled to depletion of susceptible individuals (Kermack and McKendrick, 1927; Anderson and May, 1991; Keeling and Rohani, 2008).

However, comparative analyses of respiratory epidemics have revealed substantial variation in epidemic waveform dynamics across regions. During the first COVID-19 wave, some regions exhibited sharply peaked epidemic curves with rapid post-peak decline, whereas others displayed more prolonged plateau-like dynamics over similar time intervals. Such differences in waveform structure suggest that the mechanisms governing epidemic timing and decline may vary across settings and are not readily explained by transmission feedback alone.

At the same time, it is well established that respiratory viruses are subject to seasonal and environmental modulation (Dowell, 2001; Shaman and Kohn, 2009; Tamerius et al., 2011). Factors such as temperature, humidity, ultraviolet radiation, and seasonal variation in host susceptibility may influence the timing and intensity of population-level exposure. In this perspective, epidemic dynamics may reflect not only transmission processes but also temporally structured external drivers of viral activity.

One possible interpretation is that such structured activity reflects variation in access to a broader environmental reservoir of viral material within the biosphere, although the present study does not depend on a specific mechanistic interpretation of such a reservoir.

In this study, we examine a minimal activity-driven depletion framework in which epidemic incidence arises from the interaction between a time-varying activity function and a finite vulnerable population. The activity function is formulated as a sigmoidal activation process, analogous to other biological systems exhibiting seasonal activation patterns, such as pollen release or vegetation growth. Rather than modelling infection incidence directly, the framework operates at the level of observable endpoints (hospitalizations and mortality).

The objective of the present study is to evaluate whether this externally driven formulation is sufficient to reproduce the observed large-scale temporal structure of epidemic waveforms.

## 2. Model and methods

### 2.1 Model structure

We model epidemic waveforms at the level of observed endpoints (mortality and hospitalizations) using a parsimonious depletion framework.

- *V(t)* denotes the vulnerable population (proportion of the population) that can contribute to the endpoint during the analysed wave,
- *C(t)* denotes the cumulative endpoint count (deaths or hospitalizations, expressed as proportion of the population).

The model is driven by a time-varying activity function *A(t) (day*^*-1*^), interpreted as an aggregated per-capita hazard acting on the vulnerable population.

The structural components of the SEVA framework are illustrated schematically in Figure 1.

**Figure 1.**
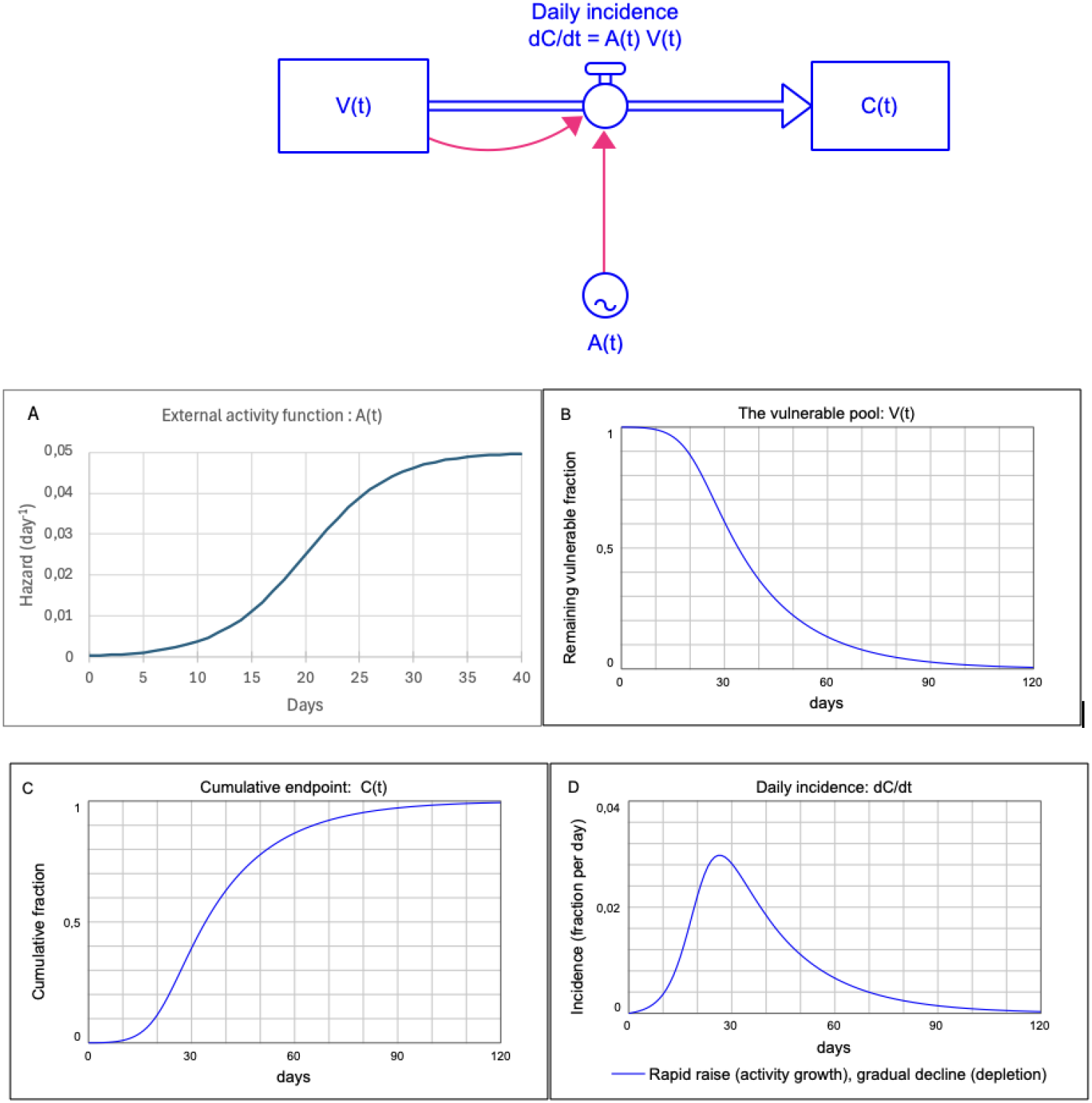
Structural dynamics of the SEVA depletion framework. Top: Conceptual structure of the model. A time-varying external activity function *A(t)* acts on a finite vulnerable population *V(t)*, generating cumulative endpoints *C(t)* with daily incidence *dC/dt = A(t)V(t)*. (A) External activity function *A(t)*. (B) Depletion of the vulnerable population *V(t)*. (C) Cumulative endpoint *C(t)*. (D) Resulting daily incidence *dC/dt*. The asymmetric rise–peak–decline waveform arises from the interaction between increasing activity and depletion of the vulnerable population.

### 2.2 Activity-driven hazard function

The activity driver is defined as:

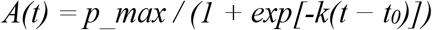

where *p_max* (day^−1^) is the maximal hazard, *k* (day^−1^) controls activation steepness, and *t*_*0*_ (days) determines the temporal midpoint of activation.

The logistic function was chosen as a minimal smooth activation profile that provides independent control of amplitude (*p_max*), timing (*t*_*0*_), and steepness (*k*).

It yields bounded growth, ensures differentiability, and avoids discontinuities that would introduce artificial kinks in the hazard profile. These properties allow transparent separation of intensity and time-scale effects in the resulting depletion dynamics.

Although asymptotic in time, for the parameter values used (*k* = 0.25, *t*_*0*_ = 20), activity increases from approximately 0.7% to 99.3% of its maximum between day 0 and day 40, effectively corresponding to a finite activation window.

### 2.3 Dynamical equations

Endpoint accumulation is described by the system:

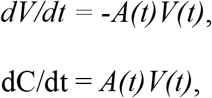

with initial conditions *V(0) = V*_*0*_ and *C(0)* = 0.

The system conserves the endpoint-relevant population:

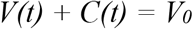

Expressed in fractional form, defining

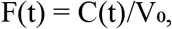

the system can be written as

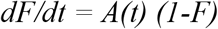

This formulation highlights that the daily incidence arises from the interaction between the activity driver and the remaining undepleted fraction of the vulnerable population.

Because activity increases monotonically while the vulnerable population is simultaneously depleted, the interaction produces an asymmetric rise–peak–decline waveform.

### 2.4 Parameter exploration

We examined the influence of the parameters *p_max, t*_*0*_, and *k* on the temporal waveform properties of the daily incidence *dC/dt*. (Fig. 2)

**Figure 2.**
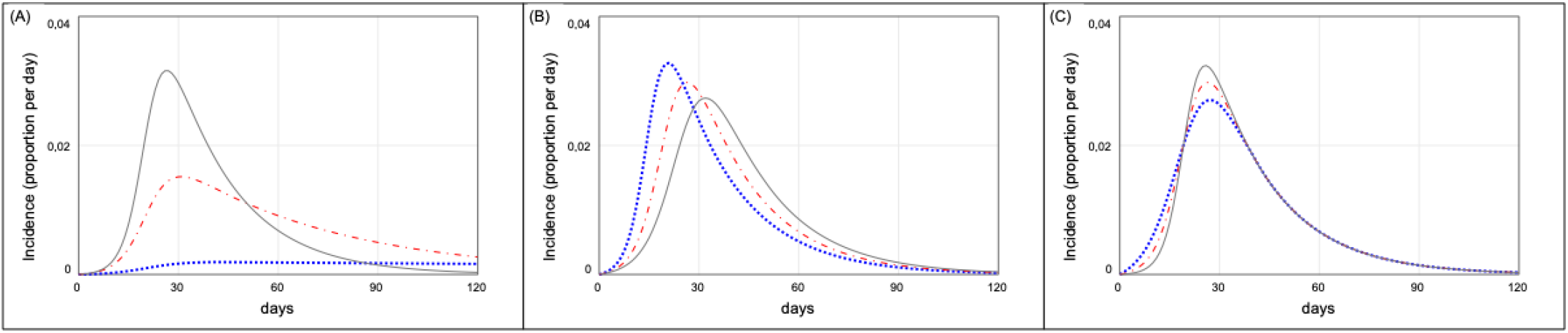
Parameter sensitivity of the activity-driven depletion model. (A) *p_max* varied (0.002, 0.02, 0.05) with fixed *t*_*0*_ = 20 and *k* = 0.25. (B) *t*_*0*_ varied (15, 20, 25) with fixed *p_max* = 0.05 and *k* = 0.25. (C) *k* varied (0.20, 0.25, 0.30) with fixed *p_max* = 0.05 and *t*_*0*_ = 20. Dotted, dashed, and solid lines represent increasing parameter values.

Parameter sensitivity analysis demonstrates distinct structural roles of the three parameters:

- *p_max* primarily controls peak amplitude and the rate of post-peak depletion, thereby influencing both the height of the peak and the duration of the declining phase.
- *t*_*0*_ shifts the waveform along the time axis with minimal effect on its intrinsic shape.
- *k* modulates the acceleration phase by controlling the steepness of activation.

These parameter roles illustrate how waveform structure arises from the interaction between activity and depletion.

This parameter dependence reflects the underlying dynamical balance between activity growth and depletion of the vulnerable population. Within the SEVA formulation the epidemic turning point occurs when the growth of the activity function is balanced by depletion of the vulnerable population.

### 2.5 Interpretation of the vulnerable population

The vulnerable population *V(t)* is endpoint-specific. For mortality, *V(t)* represents the effective subset of the population capable of contributing to deaths within the analysed wave. For hospitalization, it represents the corresponding subset capable of contributing to hospital admissions.

The size of this population implicitly captures heterogeneity in age structure, comorbidity burden, and pre-existing protection within a given region and time window. The proportionality assumption is local to the analysed wave and region.

This formulation avoids explicit modeling of infection incidence. Because infection incidence is not directly observed, parameters such as the infection fatality ratio (IFR) are not independently identifiable from endpoint data alone (Brookmeyer and Gail, 1988).

### 2.6 Data sources

#### 2.6.1 Mortality data – Europe

Observed COVID-19 mortality data for European countries were obtained from the European Centre for Disease Prevention and Control (ECDC) COVID-19 surveillance database (European Centre for Disease Prevention and Control, 2020). Daily death counts were expressed per 100,000 population and smoothed using a 7-day moving average prior to analysis.

#### 2.6.2 Mortality data – United States

State-level mortality data for the United States were obtained from the COVID Tracking Project (The COVID Tracking Project, 2021), which compiled harmonized daily death counts reported by official state and territorial health authorities. Daily mortality rates per 100,000 population were computed and smoothed using a 7-day moving average prior to analysis.

#### 2.6.3 Reporting variability and smoothing

Daily mortality reporting exhibits substantial short-term variability due to reporting delays, weekend effects, and administrative batching. The 7-day moving average therefore provides a more stable representation of the underlying epidemic waveform used for model comparison (Fig. 3).

**Figure 3.**
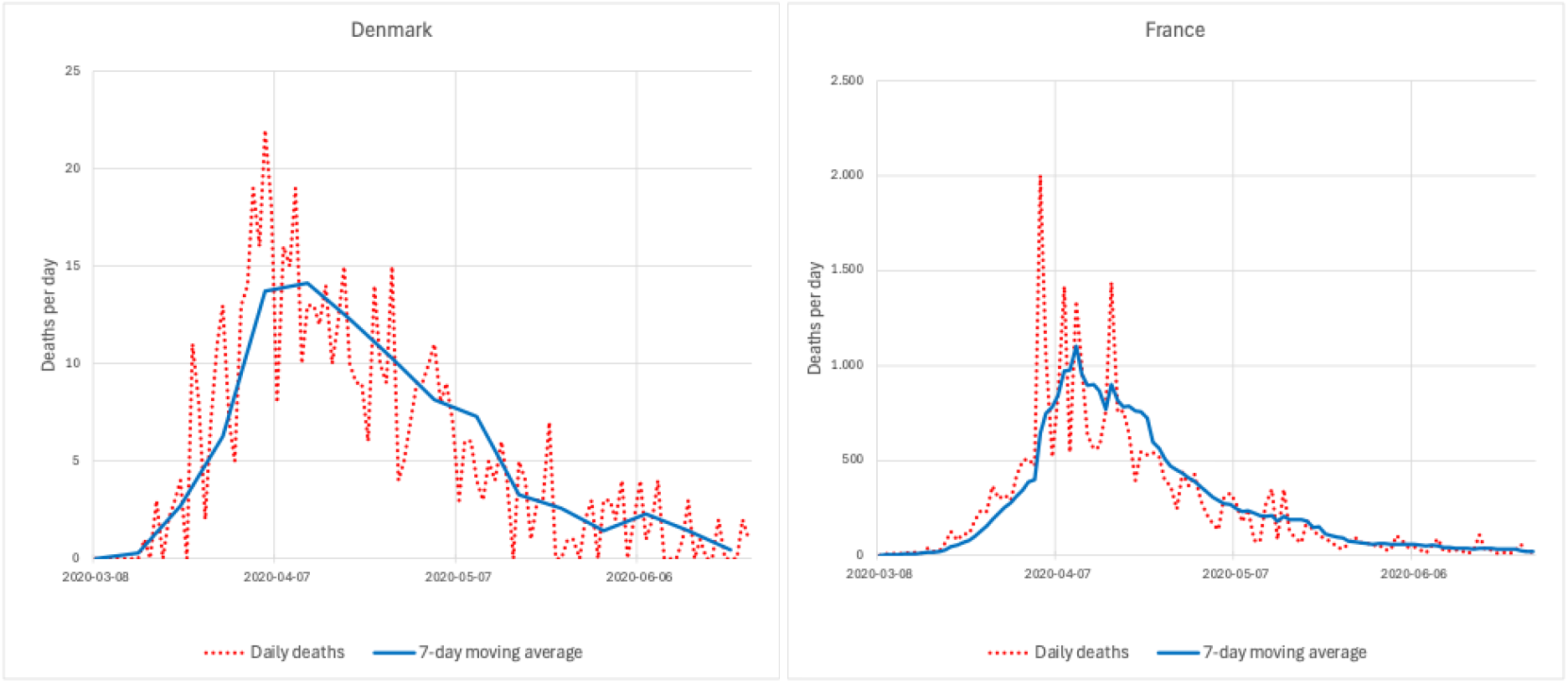
Reporting variability in daily mortality data. The figure shows daily reported COVID-19 deaths in Denmark and France from the ECDC surveillance dataset together with the corresponding 7-day moving average calculated in this study.

#### 2.6.4 Hospitalization data

Hospital admission data were obtained from *Our World in Data* (Mathieu et al., 2020), which aggregates officially reported national datasets. Weekly hospital admissions per million population were converted by Our World in Data to daily values using linear interpolation between weekly reporting points. No additional smoothing was applied to these series prior to temporal alignment and model comparison. Because the underlying data represent weekly aggregates, the interpolated daily series already exhibit reduced short-term variability relative to raw daily reporting data.

All analyses focused on the first epidemic wave during spring 2020, corresponding approximately to the first four months following epidemic onset in each region. Quantitative goodness-of-fit metrics were computed over a standardized 100-day window within this period to ensure comparability across regions.

### 2.7 Calibration procedure

Model application was performed in two sequential steps designed to separate endpoint magnitude from temporal waveform structure.

#### Default activation parameters

Unless otherwise specified, activation timing and steepness were fixed at *t*_*0*_ = 20 days and *k* = 0.25 day^−1^. Sensitivity analysis (Fig. 2) showed that waveform morphology is robust to moderate variation in these parameters. Fixing *t*_*0*_ and *k* therefore reduces parameter dimensionality while preserving the essential dynamical behaviour.

#### Step 1: Endpoint magnitude

For each region and endpoint, the initial vulnerable population V(0) was determined by matching the cumulative model output to the observed cumulative endpoint total over the analysis window. This step isolates differences in overall endpoint magnitude from differences in temporal waveform structure.

#### Step 2: Waveform structure

With endpoint magnitude fixed, the activity intensity parameter *p_max* was varied to reproduce the characteristic temporal features of the observed daily endpoint curves, including peak presence or absence, waveform asymmetry, and rate of decline.

Parameter values were selected to achieve correspondence in these structural features rather than to minimize residual error. This approach reflects the focus of the analysis on identifying and reproducing intrinsic waveform dynamics rather than performing formal curve fitting. No formal optimization procedure was applied. The resulting parameter values should therefore be interpreted as effective descriptors of waveform structure at the endpoint level rather than as independently identifiable biological parameters.

### 2.8 Temporal alignment

Because the timing of underlying infections is not directly observable, model–data comparisons were performed using temporal alignment of waveform structure.

For each region and endpoint, a single constant time shift was applied to align simulated and observed curves. The shift was determined by matching peak timing and overall waveform morphology in the daily endpoint series.

### 2.9 Goodness-of-fit metrics

Model fit was quantified using root mean square error (RMSE), coefficient of determination (R^2^), and Pearson correlation coefficient (r), computed on temporally aligned daily endpoint series.

For each region and endpoint, the analysis window was defined as a fixed 100-day interval beginning on the date of the first recorded death. This standardized window was used to ensure comparability of waveform dynamics across regions.

All metrics were calculated on daily incidence rather than cumulative counts, as cumulative series are monotonic and can artificially inflate goodness-of-fit statistics through integration of small residual errors.

### 2.10 Normalization

For cross-regional comparison of waveform morphology independent of absolute endpoint magnitude, we constructed a dimensionless representation by scaling cumulative endpoints to unity at the end of the analysis window *T*.

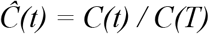

where T denotes the final day of the analysis window.

The corresponding daily incidence was expressed as a fraction of total cumulative endpoints:

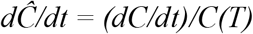

This normalization removes differences in absolute endpoint magnitude and isolates intrinsic waveform structure.

To illustrate the behaviour of the normalized representation under the SEVA formulation, Fig. 4 shows simulated normalized trajectories for a range of activity intensities *p_max* while keeping the activation parameters *k* and *t*_*0*_ constant. Under this representation, differences in absolute epidemic size are removed and the remaining variation reflects differences in activity intensity and the resulting depletion dynamics.

**Figure 4.**
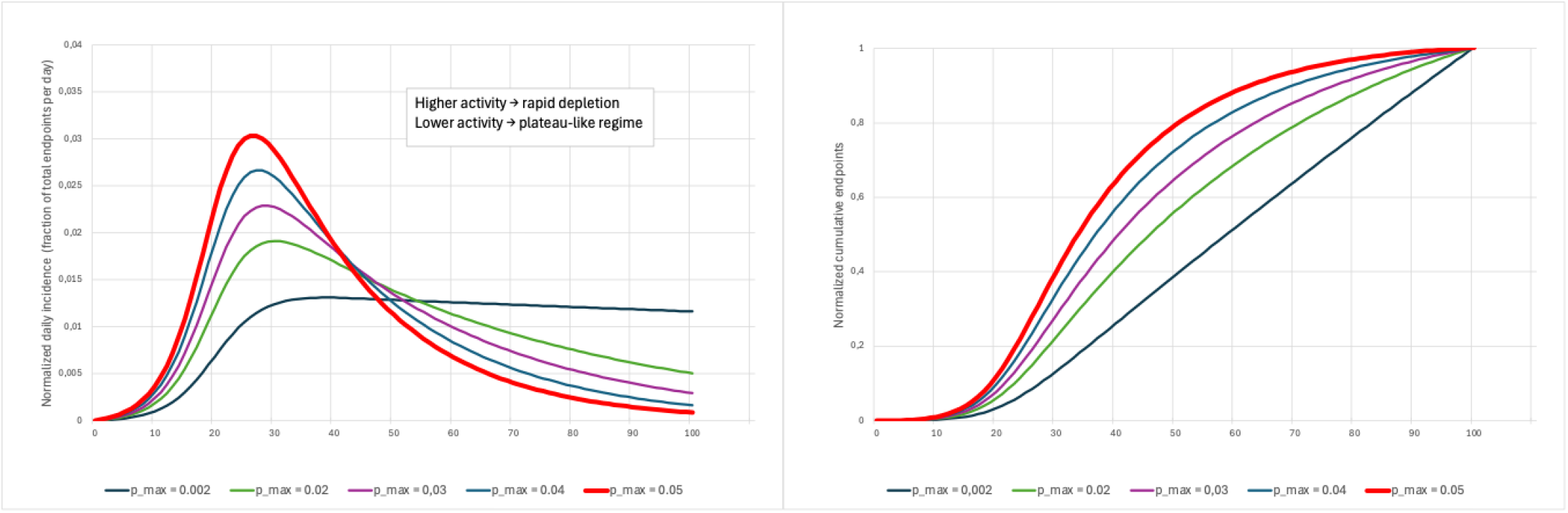
Theoretical normalized epidemic trajectories under the SEVA framework. Left: normalized daily incidence *(dC/dt)/C(T)*. Right: normalized cumulative endpoint *C(t)/C(T)*. Curves are shown for varying values of the activity intensity parameter *p_max* while keeping *k* and *t*_*0*_ constant. The normalized representation removes differences in absolute epidemic size and illustrates the waveform regimes expected under the SEVA formulation.

For the reference parameter setting used in this study (*k* = 0.25, *t*_*0*_ = 20), the simulated turning point in the high-activity regime occurs at approximately day 28. This reference behaviour provides a useful baseline for comparison with the normalized empirical curves presented in the Results section.

### 2.11 Simulation environment

All model simulations were implemented using Stella Architect (Isee Systems, Lebanon, New Hampshire, USA). The software was used to define the stock–flow structure, parameterize model equations, and generate time-series outputs for comparison with empirical endpoint data.

Simulations were performed using explicit Euler integration with a daily time step (Δt = 1 day). Given the characteristic time scale of activation (k ≈ 0.25 day^−1^), this discretization is sufficient to ensure numerical stability and negligible integration error relative to waveform morphology.

In the graphical implementation, the activation window was specified over a 40-day interval. This corresponds to the analytical logistic formulation with k = 0.25 day^−1^ and t_0_ = 20 days, for which activity increases from approximately 0.7% to 99.3% of its maximum between day 0 and day 40.

## 3. Results

### 3.1 Separation of epidemic waveform dynamics and endpoint magnitude

A direct comparison between regions with markedly different mortality burdens illustrates a separation between epidemic waveform dynamics and absolute endpoint magnitude (Fig. 5). New York State and Norway represent contrasting cases, with New York experiencing substantially higher cumulative mortality during the first epidemic wave.

**Figure 5.**
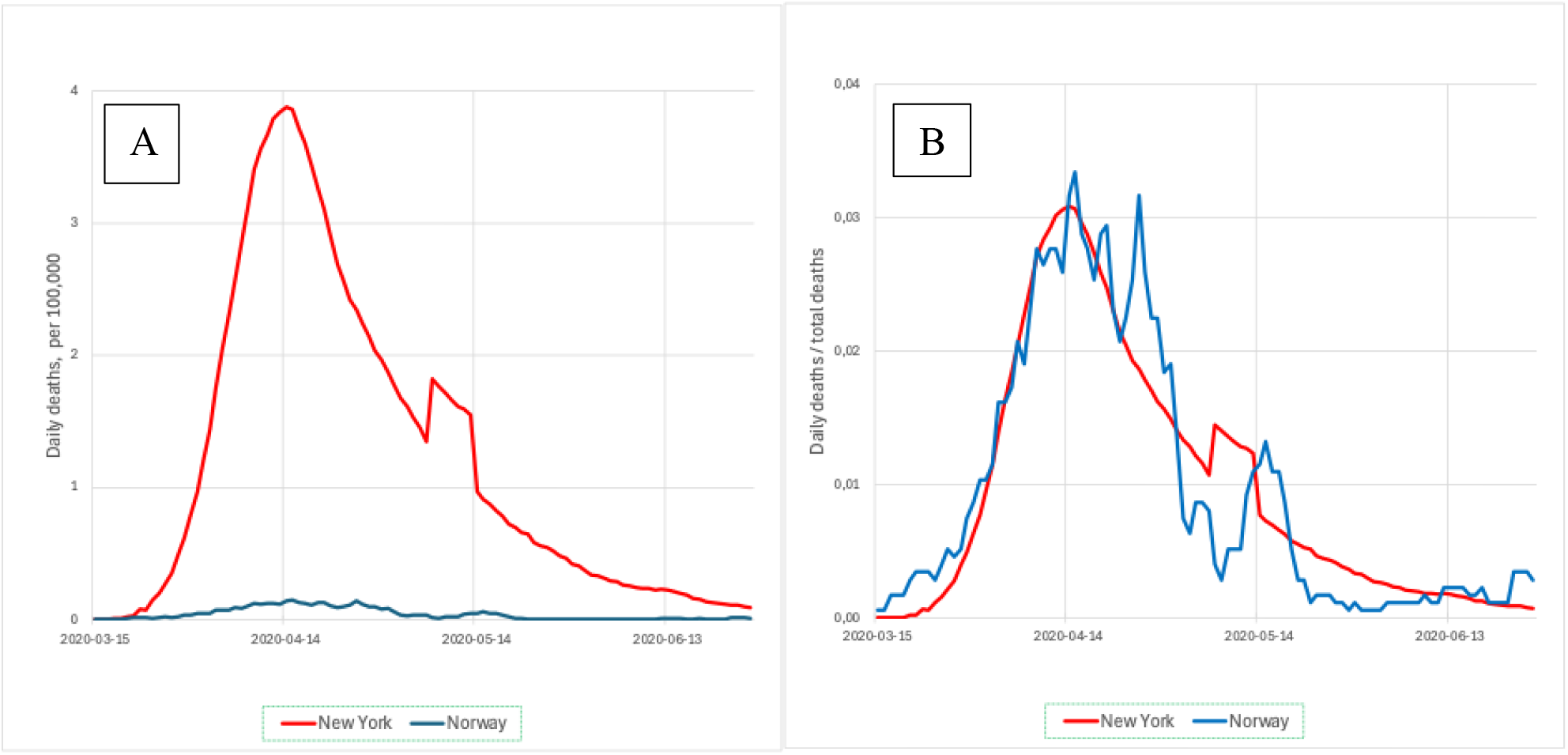
Separation of epidemic waveform dynamics and mortality magnitude in New York and Norway. Despite this large difference in absolute mortality, the normalized daily mortality curves display highly similar temporal structure. Both regions exhibit a rapid rise, a well-defined peak, and a subsequent decline occurring over a comparable time scale. The timing of the turning point and the overall waveform asymmetry are closely aligned after normalization.

These observations indicate that epidemic waveform dynamics can be largely independent of absolute endpoint magnitude. While cumulative mortality reflects the size of the endpoint-specific vulnerable population, the temporal evolution of the epidemic curve appears to follow a common structural pattern across regions.

### 3.2 Normalized waveform regimes

To examine waveform dynamics independent of absolute magnitude, cumulative mortality was normalized to unity at the end of the analysis window, and daily mortality was expressed as a fraction of total cumulative mortality (see Section 2.10).

Under this dimensionless representation, two distinct classes of epidemic waveform behaviour are observed across regions (Fig. 6).

**Figure 6.**
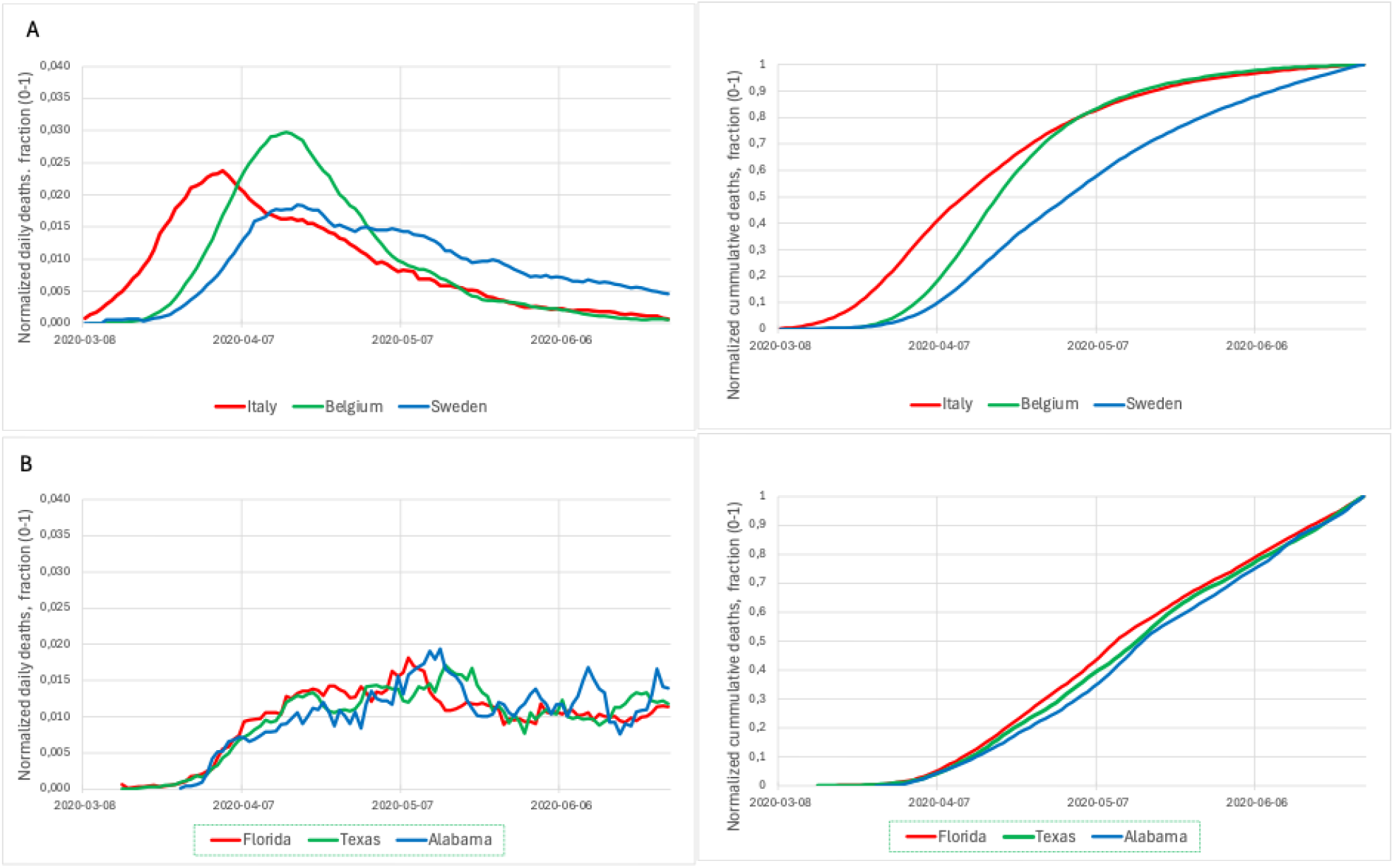
Empirical normalized epidemic waveform regimes. Normalized cumulative and daily COVID-19 mortality curves are shown for representative regions during the first epidemic wave. Cumulative mortality is scaled to unity at the end of the analysis window, and daily mortality is expressed as a fraction of total cumulative mortality. Panel A shows regions exhibiting a turning-point regime, characterized by a distinct peak followed by sustained decline and a sigmoidal cumulative trajectory. Panel B shows regions exhibiting an activation-dominated regime, in which daily incidence remains elevated without a pronounced decline and cumulative mortality displays near-linear growth.

In the first class, a clear turning point occurs within the analysis window, followed by a sustained decline in normalized daily incidence (Fig. 6A). These curves exhibit a pronounced unimodal structure and corresponding sigmoidal cumulative trajectories, consistent with substantial depletion of the endpoint-specific vulnerable population during the observed period.

In the second class, no pronounced turning point is observed within the analysis window (Fig. 6B). Normalized daily incidence rises and remains elevated without sustained decline, while cumulative mortality displays near-linear growth over much of the interval. This pattern is consistent with limited depletion relative to ongoing activation within the observation window.

These two waveform regimes provide an empirical classification of epidemic dynamics across regions. The distinction is based on intrinsic temporal structure rather than absolute mortality magnitude and forms the basis for subsequent comparison with model-derived dynamics.

### 3.3 Model-based interpretation of waveform regimes

The waveform regimes identified above can be interpreted within the SEVA framework as arising from the interaction between a time-varying activity driver and depletion of a finite vulnerable population.

Using fixed activation parameters (*t*_*0*_ = 20, *k* = 0.25), the model reproduces both classes of waveform behaviour through variation in the activity intensity parameter *p_max*. For higher values of *p_max*, the activity-driven hazard rapidly depletes the vulnerable population, resulting in a pronounced peak followed by sustained decline within the observation window, corresponding to the turning-point regime.

For lower values of *p_max*, depletion proceeds more slowly relative to the rate of activation. As a result, daily incidence remains elevated without a clear turning point within the same time window, corresponding to the activation-dominated regime.

Within this formulation, the presence or absence of a turning point reflects the balance between activity intensity and depletion of the vulnerable population. The model therefore provides a unified dynamical interpretation of the two empirically identified waveform regimes without requiring changes to the underlying model structure.

These relationships are illustrated in the normalized model trajectories (Fig. 4), which show that the transition between peaked and plateau-like dynamics emerges continuously as a function of activity intensity under fixed activation timing and steepness.

The extent of depletion within the observation window can be quantified by the depletion fraction *C(T)/V*_*0*_, representing the proportion of the endpoint-specific vulnerable population depleted by time *T*.

For low activity intensity (*p_max* = 0.002) with fixed activation parameters (*t*_*0*_ = 20, *k* = 0.25), the model yields *C(T)/V*_*0*_ ≈ 0.18 at *T* = 26 June 2020. This indicates that the majority of the vulnerable population remains undepleted within the first-wave window, consistent with the activation-dominated regime.

In contrast, higher activity intensity leads to substantially greater depletion within the same time window, resulting in the emergence of a turning point and subsequent decline in daily incidence.

### 3.4 Regional mortality dynamics in the United States

Figure 7 contrasts mortality dynamics between selected northern and southern U.S. states during the first epidemic wave. The analysis window was restricted to the initial epidemic period (through 5 July 2020) in order to isolate first-wave dynamics prior to subsequent summer increases.

**Figure 7.**
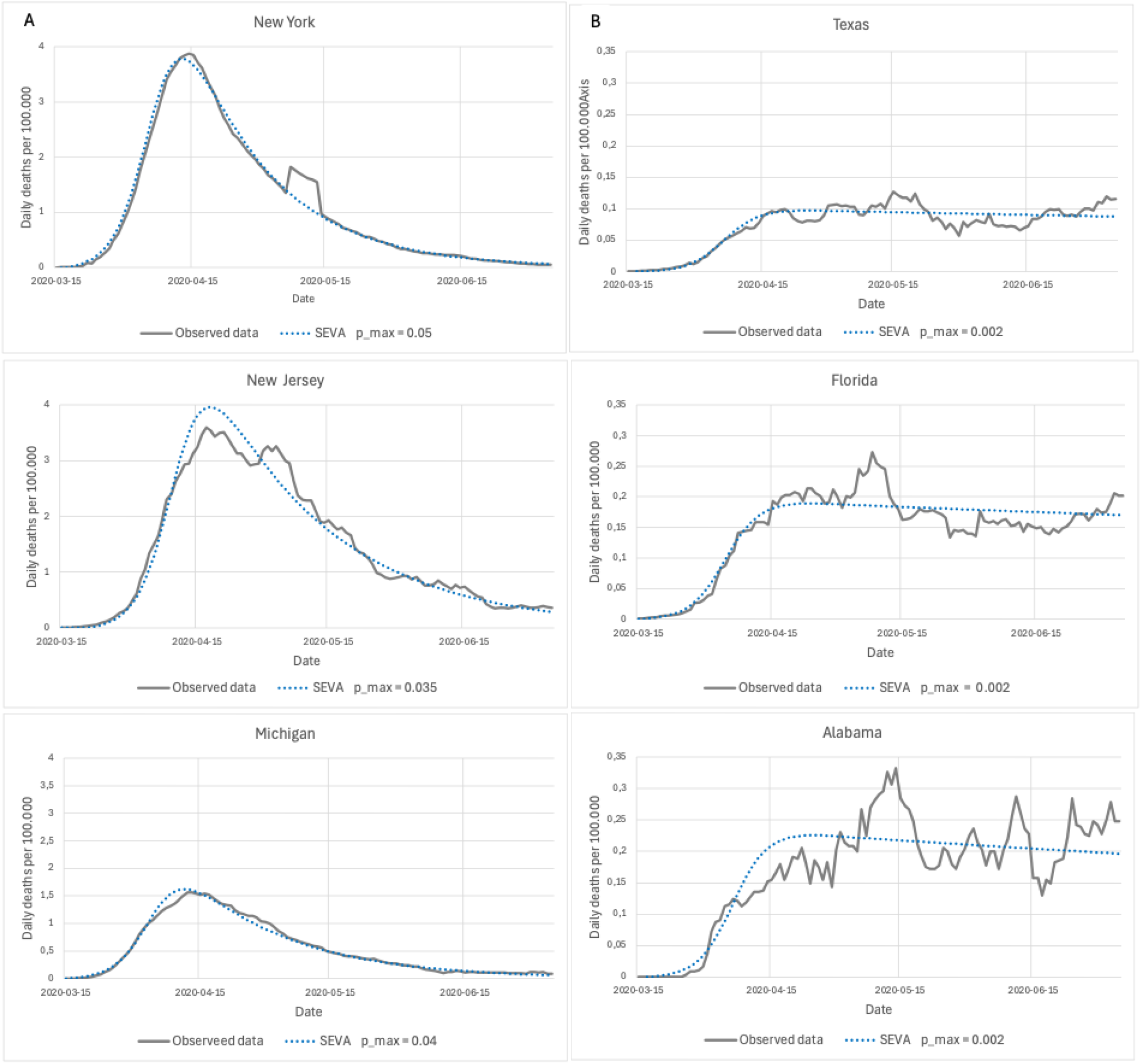
Regional contrast in U.S. mortality dynamics. Observed daily COVID-19 mortality (per 100,000 population) is shown together with SEVA model outputs for selected northern (Panel A) and southern (Panel B) U.S. states during the first epidemic wave. Model simulations use fixed activation parameters (t_0_ = 20, k = 0.25). Note: y-axis scales differ between panels.

The contrast between northern and southern states illustrates two distinct dynamical regimes within the SEVA framework. With activation timing and steepness held constant, both waveform types are reproduced through variation in activity intensity and endpoint scaling.

In northern states, the turning point occurs within the analysis window and corresponds to substantial depletion of the endpoint-specific vulnerable population. In southern states, the absence of sustained decline is consistent with incomplete depletion under lower activity intensity.

These observations are consistent with the interpretation that diverse epidemic waveform morphologies across regions can be reproduced within the same activity-driven depletion framework.

### 3.5 Mortality dynamics in European countries

To examine whether the SEVA framework can reproduce the empirically observed waveform structures, the model was applied to mortality data from a set of European countries during the first epidemic wave (Fig. 8). The same countries are analysed in the corresponding hospitalization analysis (Fig. 9), allowing direct comparison across endpoints.

**Figure 8.**
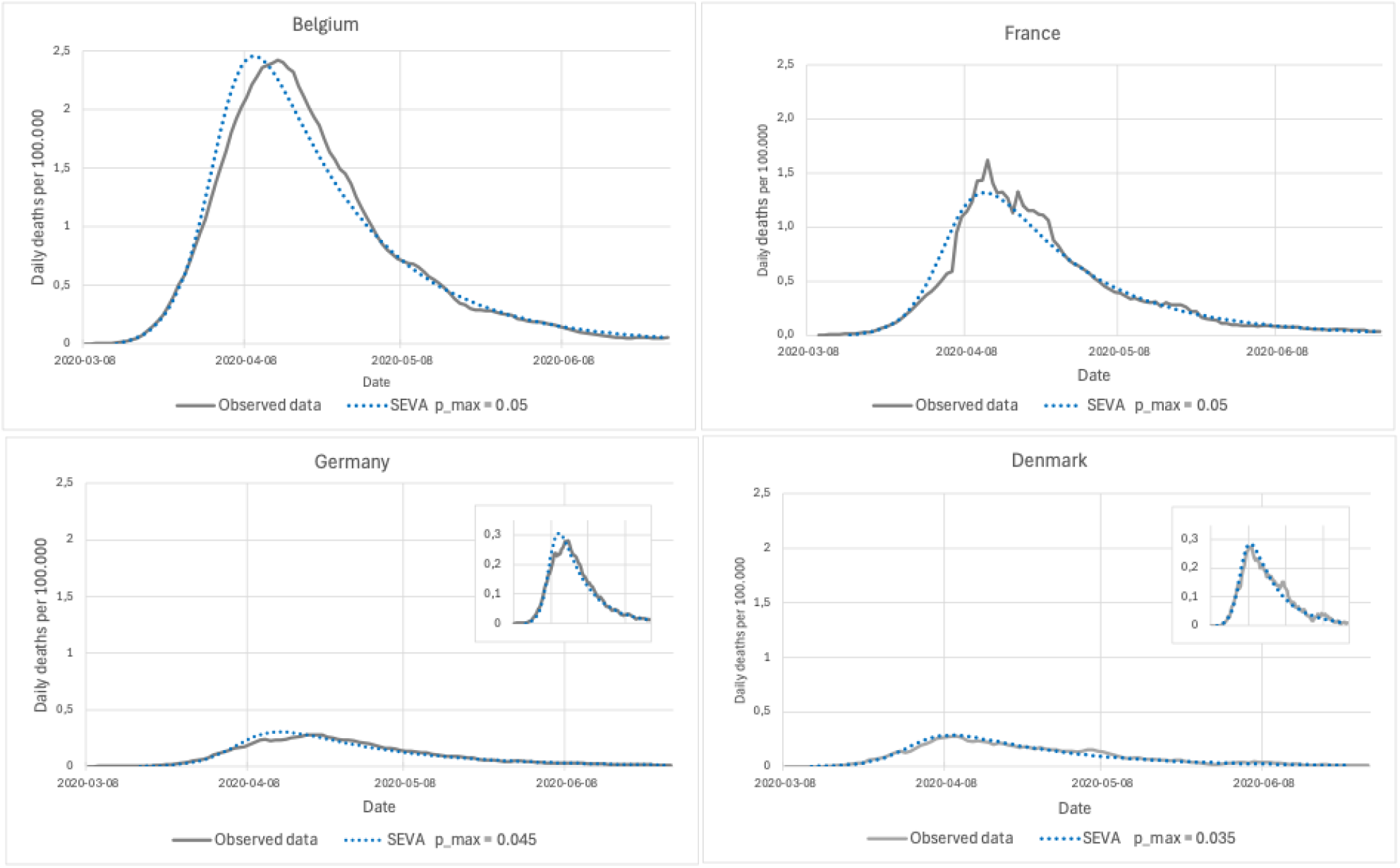
Mortality incidence during the first COVID-19 wave in selected European countries. Observed daily COVID-19 mortality (per 100,000 population) is shown together with SEVA model outputs using fixed activation parameters (*t*_*0*_ = 20, *k* = 0.25) and country-specific *p_max*. Insets in the Germany and Denmark panels show the same data on a rescaled y-axis to highlight waveform structure in lower-incidence settings.

**Figure 9.**
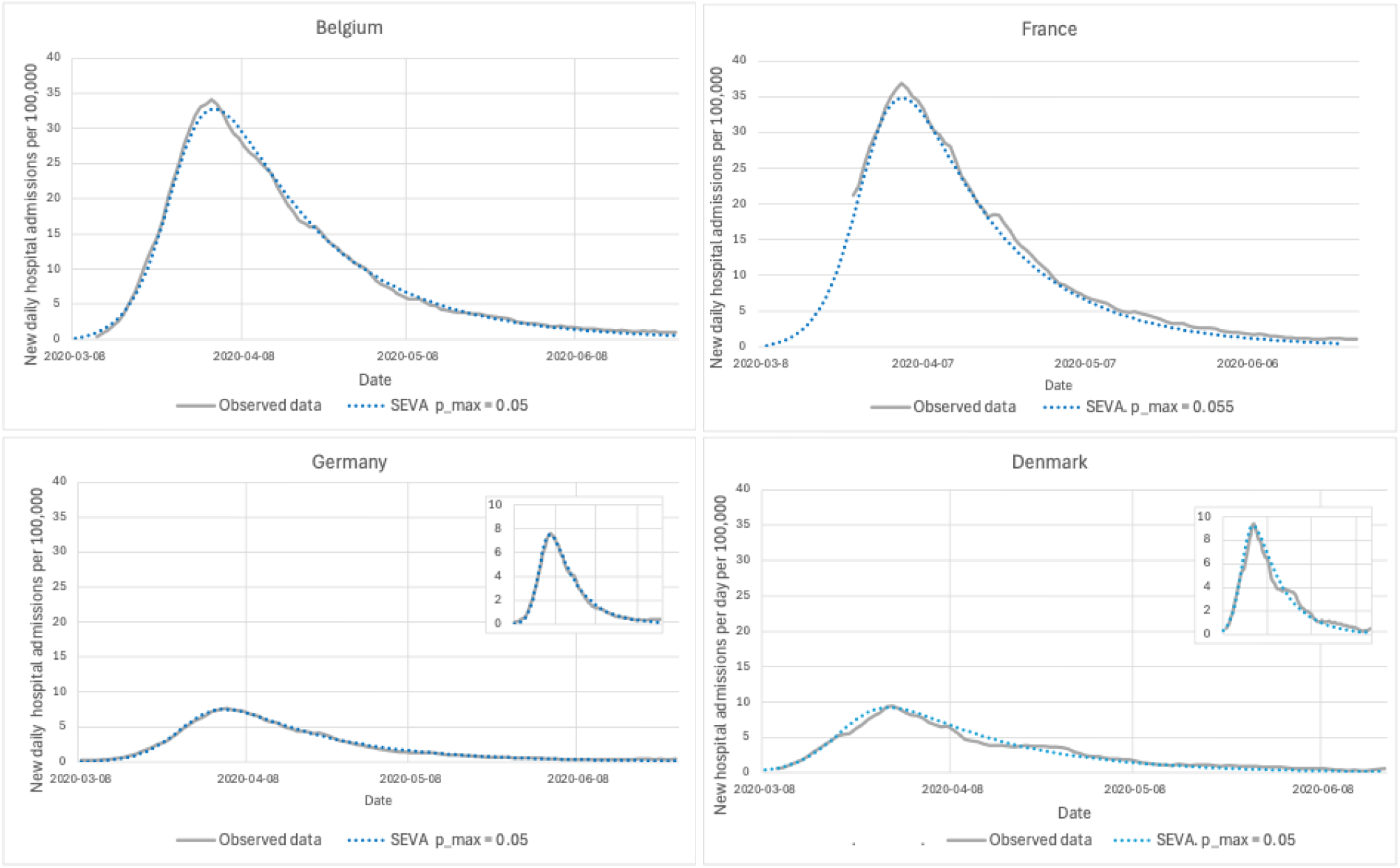
Hospitalization incidence during the first COVID-19 wave in selected European countries. Observed daily hospital admissions (per 100,000 population) are shown together with SEVA model outputs using fixed activation parameters (*t*_*0*_ = 20, *k* = 0.25) and country-specific *p_max*. Insets in the Germany and Denmark panels show the same data on a rescaled y-axis to highlight waveform structure in lower-incidence settings.

Across countries, mortality time series exhibit the characteristic waveform morphology identified above, including a rapid rise, a distinct peak, and a prolonged decline. Within the SEVA framework, these features are reproduced using a common activation profile, with differences in waveform shape accounted for by variation in activity intensity and endpoint scaling.

Inter-country differences in peak sharpness and rate of decline correspond to differences in activity intensity, while cumulative mortality levels are matched independently through scaling of the endpoint-specific vulnerable population.

Similar correspondence between observed and simulated waveforms is observed across 12 additional European countries (Fig. S1), supporting the general applicability of the activity-driven depletion framework across diverse epidemiological settings.

### 3.6 Hospitalization dynamics in European countries

To assess whether the identified waveform structures extend across clinical endpoints, the SEVA framework was applied to hospitalization data from the same set of European countries analysed above (Fig. 9).

Hospitalization time series exhibit the same characteristic waveform morphology as observed for mortality, including a rapid rise, a distinct peak, and a more gradual decline, despite representing a different clinical endpoint.

Within the SEVA framework, these dynamics are reproduced using the same activation parameters, with differences in waveform shape accounted for by variation in activity intensity and endpoint scaling.

The consistency of waveform structure across mortality and hospitalization data supports the interpretation that the observed epidemic dynamics reflect a common underlying process, with endpoint-specific differences primarily affecting magnitude rather than temporal form.

### 3.7 Quantitative fit performance

Goodness-of-fit was evaluated on temporally aligned daily mortality incidence using RMSE, MAE, R^2^, and Pearson correlation (r) over a standardized 100-day window defined as the first 100 days following the initial recorded death in each region (Table 1).

**Table 1.** Quantitative agreement between observed and simulated COVID-19 mortality curves. Goodness-of-fit metrics (RMSE, MAE, R^2^) were calculated on aligned daily mortality series over a fixed 100-day analysis window. Metrics reflect correspondence in temporal waveform morphology following independent matching of cumulative endpoint magnitude.

| Region | $p_{max}$ | RMSE | MAE | $R^2$ | Pearson_r |
| --- | --- | --- | --- | --- | --- |
| Belgium | 0.050 | 0.14 | 0.09 | 0.97 | 0.98 |
| Netherlands | 0.045 | 0.07 | 0.05 | 0.95 | 0.98 |
| Spain | 0.050 | 0.15 | 0.10 | 0.94 | 0.97 |
| UK | 0.035 | 0.05 | 0.04 | 0.99 | 0.99 |
| Michigan | 0.040 | 0.06 | 0.04 | 0.98 | 0.99 |
| New Jersey | 0.035 | 0.24 | 0.18 | 0.96 | 0.98 |
| New York State | 0.050 | 0.23 | 0.14 | 0.96 | 0.98 |
| Alabama | 0.002 | 0.04 | 0.04 | 0.59 | 0.77 |
| Florida | 0.002 | 0.02 | 0.02 | 0.88 | 0.94 |
| Texas | 0.002 | 0.01 | 0.01 | 0.86 | 0.93 |

Because cumulative endpoint magnitude was matched independently during calibration, the reported metrics primarily quantify correspondence in temporal waveform morphology, including peak timing, amplitude, and decline structure - rather than differences in total mortality burden.

Across European countries and U.S. states, the metrics indicate close structural correspondence between simulated and observed daily mortality curves under the unified SEVA framework.

## 4. Discussion

The present study examines whether the large-scale waveform structure observed during the first COVID-19 epidemic wave can be reproduced within a minimal activity-driven depletion framework. The results show that a simple formulation in which a time-varying activity function acts on a finite vulnerable population is sufficient to generate the characteristic asymmetric rise–peak–decline patterns observed in hospitalization and mortality data across multiple regions.

While interpersonal transmission undoubtedly occurs, the observed epidemic waveforms during the first COVID-19 wave are not readily explained by transmission feedback alone. The present results show that the large-scale temporal structure of these waveforms can be reproduced without invoking transmission feedback as the primary dynamical driver. In this perspective, the temporal evolution of epidemic incidence emerges from the interaction between externally structured activity and depletion of a finite vulnerable population.

Within the SEVA framework, epidemic incidence arises from the interaction between a temporally structured activity function A(t) and depletion of the vulnerable population V(t). As activity increases over time, incidence initially rises; as the vulnerable population becomes progressively depleted, incidence subsequently declines. The epidemic turning point occurs when the increase in activity is balanced by depletion of the remaining vulnerable population. This mechanism produces asymmetric epidemic waves without requiring explicit modelling of transmission feedback or infection-to-event delay distributions.

The model further indicates that distinct epidemic waveform regimes may arise under different activity intensities. Higher activity intensity leads to rapid depletion of the vulnerable population and produces sharply peaked epidemic waves with sustained post-peak decline. In contrast, lower activity intensity results in incomplete depletion within the observation window, yielding plateau-like incidence trajectories. These two dynamical regimes correspond to contrasting epidemic patterns observed during the first COVID-19 wave.

A central empirical observation is the marked similarity of normalized epidemic waveforms across regions with very different absolute mortality burdens. When cumulative mortality and daily incidence are expressed relative to the final epidemic size, curves from geographically and demographically distinct regions display closely similar temporal structures. Within the SEVA formulation, this behaviour arises because normalized trajectories depend primarily on the temporal structure of the activity function rather than on the absolute size of the vulnerable population. If the temporal profile of viral activity is broadly similar across regions, normalized epidemic waveforms will tend to converge toward similar shapes.

Mathematically, the SEVA formulation is closely related to hazard-based models widely used in survival analysis. In this interpretation, the activity function A(t) represents a time-varying population hazard acting on the remaining vulnerable fraction of the population. Epidemic incidence can therefore be interpreted as the product of exposure intensity and the remaining population capable of contributing to observable endpoints, while the cumulative trajectory reflects the integrated activity acting on the vulnerable population over time.

Epidemic curves during the first COVID-19 wave have frequently been described as approximately Gompertz-like. In transmission-based epidemic models, such behaviour has often been interpreted as evidence of strong heterogeneity in transmission or superspreading dynamics. However, Gompertz-type trajectories can also be expressed within a hazard–depletion formulation mathematically similar to the SEVA model. The present results therefore indicate that Gompertz-like epidemic waveforms do not uniquely imply transmission-driven dynamics, but may also emerge from activity-driven hazard processes in which transmission feedback is not the primary determinant of temporal structure.

Within the SEVA framework, the activity function should be interpreted as an aggregate representation of temporally structured viral exposure rather than as a directly measurable environmental variable. Multiple biological processes may contribute to such structured activity, including environmental factors such as temperature, humidity, ultraviolet radiation, and seasonal variation in host susceptibility.

At the level of large populations such as countries or states, the activity function should therefore be understood as a population-level average of highly heterogeneous local exposure processes. Viral exposure varies substantially across individuals and locations, for example when susceptible individuals come into close proximity with infected persons, where local viral load may be elevated. The SEVA formulation does not assume homogeneous exposure but instead describes the aggregate effect of heterogeneous exposure conditions acting at the population level.

More generally, sigmoidal activation followed by sustained activity is a common feature of many biological systems. Comparable activation dynamics are observed in processes such as seasonal pollen release and fungal spore dispersal. The activity function used here can therefore be viewed as a generic dynamical representation of temporally structured biological activation rather than as a claim about a single identifiable environmental mechanism.

One possible interpretation is that the activity function reflects access to a broader environmental reservoir of viral material within the biosphere. However, the present model does not identify or quantify such a reservoir and does not depend on a specific mechanistic interpretation.

The present study does not attempt to identify the specific biological origin of the activity function, nor does it attempt to reconstruct infection incidence or estimate epidemiological parameters such as reproduction numbers. Instead, the objective is to examine whether the large-scale temporal structure of epidemic waves can be reproduced by a minimal dynamical formulation operating directly at the level of observable endpoints.

Several limitations of this structural approach should be acknowledged. First, the model operates at the level of hospitalization and mortality rather than explicitly modelling infection incidence, and infection-to-event delay distributions are therefore not represented explicitly. Second, the activity function is introduced phenomenologically and is not derived from independently measured environmental variables. Third, parameter selection focuses on reproducing waveform morphology rather than formal statistical inference. The purpose of the present analysis is therefore structural comparison between model dynamics and observed epidemic waveforms rather than detailed epidemiological parameter estimation.

Despite these limitations, the structural properties of the SEVA framework provide a consistent and parsimonious explanation of the observed epidemic waveform dynamics.

## Concluding remarks

Taken together, the results indicate that large-scale epidemic waveforms can be reproduced by a simple dynamical mechanism arising from the interaction between a temporally structured activity function and depletion of a finite vulnerable population. Within this formulation, epidemic incidence reflects the product of exposure intensity and the remaining population capable of contributing to observable endpoints.

As activity increases, incidence rises; as the vulnerable population becomes progressively depleted, incidence declines, producing the characteristic asymmetric epidemic waveform. When epidemic trajectories are expressed in normalized form, their temporal structure depends primarily on the activity profile rather than on absolute endpoint magnitude. This provides a possible explanation for the observed similarity of normalized epidemic waveforms across regions with markedly different mortality burdens.

The SEVA framework therefore suggests that diverse epidemic waveform regimes—including sharply peaked and plateau-like dynamics—may arise from differences in activity intensity within a common underlying dynamical structure. In this sense, the framework can be viewed as a testable hypothesis for the dynamical origin of epidemic waveforms, which may be evaluated using data from different pathogens and epidemiological settings.

## Data availability

The datasets generated and analysed during the current study are available in the Zenodo repository: https://doi.org/10.5281/zenodo.19021158

## Declaration of generative AI and AI-assisted technologies in the manuscript preparation process

During the preparation of this work the author used ChatGPT to assist with language editing, grammer refinement, and improvement of sentence clarity. After using this tool, the author reviewed and edited the content as needed and takes full responsibility for the content of the published article.

## Notes

### Competing Interest Statement

The authors have declared no competing interest.

### Funding Statement

The author(s) received no specific funding for this work.

### Summary of Updates

Results section updated. Figure 6,8 and 9 have been updated

