## Supplemental Fig. 1 for "SEVA: An externally driven framework for reproducing COVID-19 mortality waves without transmission feedback"

S2 Fig.  SEVA model fits to daily COVID-19 mortality (7-day average, per 100,000) for 12 additional European countries during the first epidemic wave.
Curves are aligned to 8 March 2020. Variation in waveform morphology is accommodated through adjustment of the activity intensity parameter p_max.

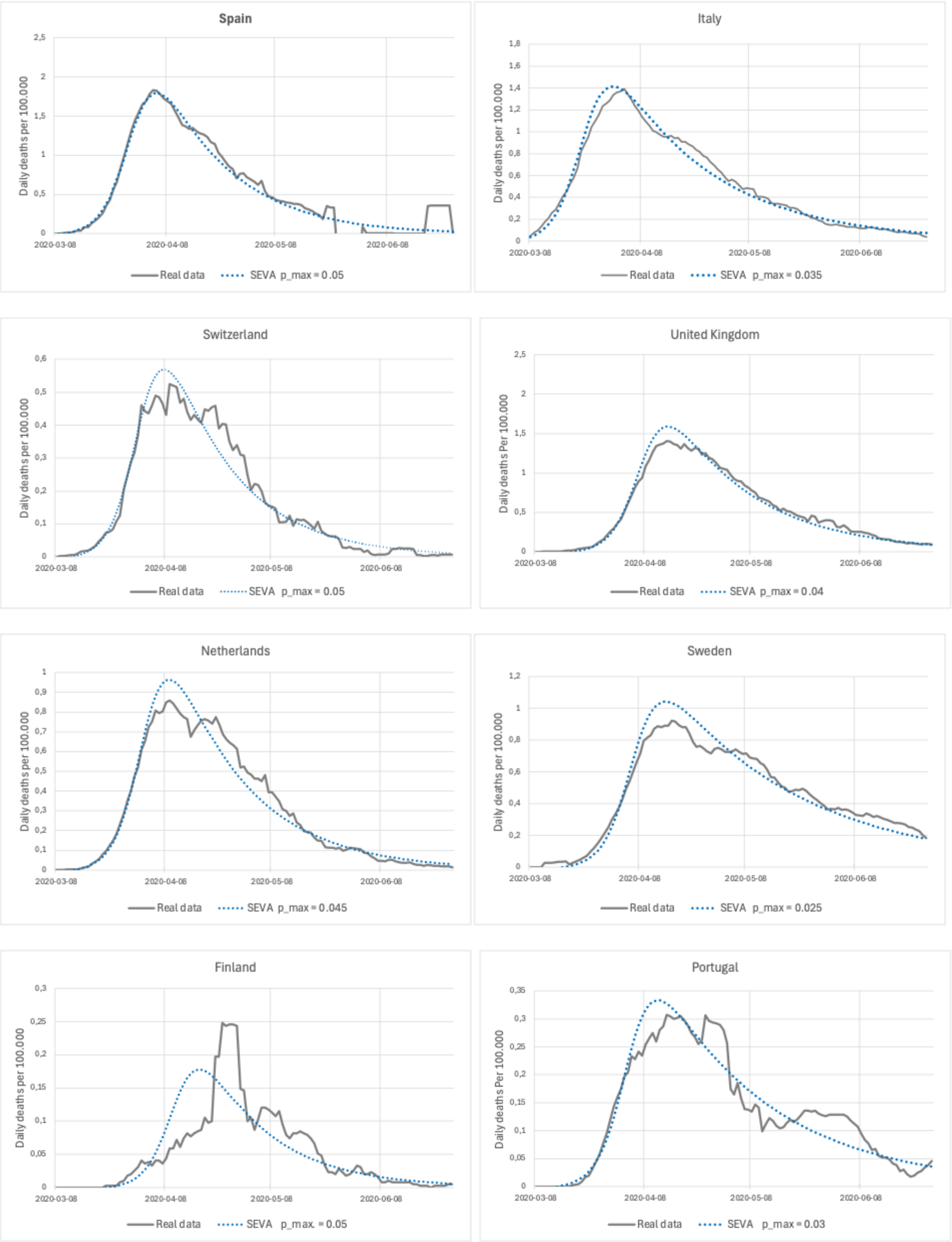

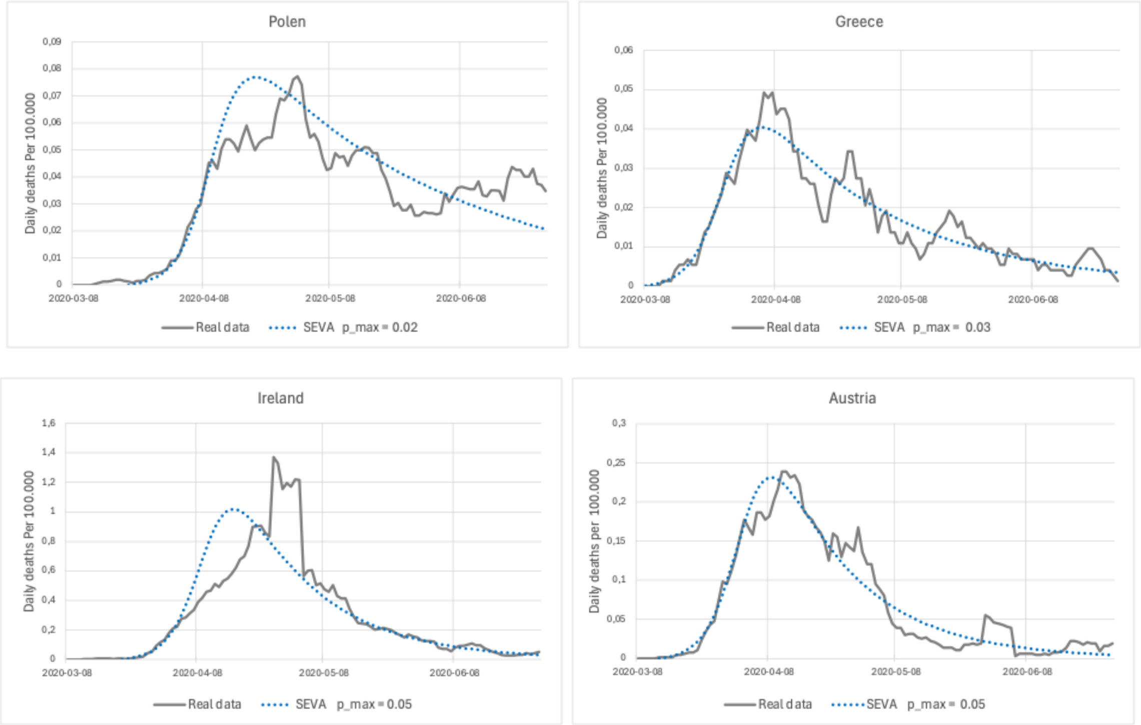

Polen
Daily deaths Per 100.000
0,09
0,08
0,07
0,06
0,05
0,04
0,03
0,02
0,01
0
2020-03-08
2020-04-08
2020-05-08
2020-06-08
Real data
SEVA p_max = 0.02
Greece
Daily deaths Per 100.000
0,06
0,05
0,04
0,03
0,02
0,01
0
2020-03-08
2020-04-08
2020-05-08
2020-06-08
Real data
SEVA p_max = 0.03
Ireland
Daily deaths Per 100.000
1,6
1,4
1,2
1
0,8
0,6
0,4
0,2
0
2020-03-08
2020-04-08
2020-05-08
2020-06-08
Real data
SEVA p_max = 0.05
Austria
Daily deaths per 100.000
0,3
0,25
0,2
0,15
0,1
0,05
0
2020-03-08
2020-04-08
2020-05-08
2020-06-08
Real data
SEVA p_max = 0.05
